# Cohort Profile: ReCoDID Consortium’s Harmonized Acute Febrile Illness Arbovirus Meta-Cohort

**DOI:** 10.1101/2023.10.10.23296846

**Authors:** Gustavo Gómez, Heather Hufstedler, Carlos Montenegro, Yannik Roell, Anyela Lozano, Adriana Tami, Tereza Magalhaes, Ernesto T.A. Marques, Angel Balmaseda, Guilherme Calvet, Eva Harris, Patricia Brasil, Victor Herrera, Luis Villar, Lauren Maxwell, Thomas Jaenisch, ReCoDID Arbovirus harmonization study group

## Abstract

Infectious disease (ID) cohorts are key to advancing public health surveillance, public policies and pandemic responses. Unfortunately, ID cohorts often lack funding to store and share clinical-epidemiological data (CE) and high-dimensional laboratory (HDL) data long-term, which is evident when the link between these data elements is not kept up to date. This becomes particularly apparent when smaller cohorts fail to successfully address the initial scientific objectives due to limited case numbers, which also limits the potential of pooling for these studies to monitor long-term cross-disease interactions within and across populations. To facilitate advancements in cross-population inference and reuse of cohort data, the European Commission (EC) and the Canadian Institutes of Health Research, Institute of Genetics (CIHR-IG) funded the ReCoDID (Reconciliation of Cohort Data for Infectious Diseases) Consortium to store and share harmonized and standardized CE and HDL data on a federated platform and also provide innovative statistical tools to conduct meta-analyses of the individual patient data. Here we describe the harmonization of CE data from nine arbovirus (arthropod-borne viruses) cohorts in Latin America, which serve as a starting point for the ReCoDID meta-cohort. CE data was retrospectively harmonized using Maelstrom’s methodology and standardized to Clinical Data Interchange Standards Consortium (CDISC).

This meta-cohort will facilitate various joint research projects, e.g., on immunological interactions between sequential flavivirus infections and for the evaluation of potential biomarkers for severe arboviral disease.

## Introduction

The meta-cohort consists of participant-level data and descriptive metadata from nine studies from five countries (Brazil, Colombia, El Salvador, Nicaragua, and Venezuela). All studies were established to study arbovirus (arthropod-borne viruses) infections in the population, with some cohorts enrolling maternal-infant pairs, or either children or pregnant women.

Most studies did not recruit participants through study-initiated contacts (e.g., emails, calls, letters, etc.). Instead, the vast majority of participants in each cohort were directly referred to the study from the health unit where they were seeking care. A small fraction of participants contacted the cohorts directly to participate because participation was associated with access to more routine care or additional screenings, which can be seen as a benefit in resource-poor settings.

The interactions between immune responses caused by different patterns of exposure over time to the four dengue virus serotypes (DENV 1-4) and Zika virus (ZIKV) have attracted considerable attention - for example as a mechanism to explain the heterogeneity in severe dengue but also in the severe outcomes seen during and after the ZIKV epidemic in Latin America(1–5). The investigation of these interactions between closely related members of the *flaviviridae* requires large sample sizes and the inclusion of populations with exposures to different sequences of pathogens, resulting in heterogeneous immune profiles.

The ReCoDID Consortium, funded by the EC and CIHR-IG(6), aims to provide ID researchers with harmonized participant-level data and metadata resources as well as analysis tools to facilitate pooled analysis projects, i.e. to advance our knowledge on the effects of prior exposure on the immune response to subsequent epidemics at the population and individual level and to inform personalized medicine approaches to diagnosis and treatment of infections. To facilitate cross-study inference in the context of emerging IDs, ReCoDID researchers created a platform to extract individual-level CE and HDL data from existing cohorts, and harmonize this data according to a specific standard. ReCoDID focuses current harmonization efforts on arbovirus and SARS-CoV- 2 cohort data, but hopes to expand these services to other IDs in the future. This cohort profile provides an overview of the newly-created arbovirus meta-cohort from five countries: Brazil, Colombia, El Salvador, Nicaragua, and Venezuela. Acute and post-acute samples were collected from each study. Information extracted from samples vary from study to study and include DENV molecular tests, DENV serotype, DENV viral load, ZIKV molecular test,, ZIKV viral load, CHIKV molecular tests and CHIKV viral load. Height, weight, birthdate, negative health outcomes associated with severe dengue (such as occurrence of bleeding, e.g.) and required interventions have also been collected. The ReCoDID Consortium aims to build a data sharing platform for linking CE data to HDL data (e.g., human and pathogen genomic data, human metabolomic and immunomics data) at the participant level that are collected from ID focused cohorts. While ReCoDID is working to share data related to other disease types, this paper describes the acute febrile illness (AFI) meta-cohort that includes, at the date of publishing this article, data from nine studies that have committed to sharing CE and HDL data from their arbovirus cohorts. All participating cohorts have submitted genomic sequences of the DENV virus to ReCoDID, except for the cohorts in Nicaragua and the Cohort of Symptomatic Pregnant Women in Brazil. Two studies— PHBDC and IDAMS cohorts— have also agreed to share genomic sequences for CHIKV and ZIKV viruses. Most participating cohorts collected and stored blood samples; Nicaragua also collected urine and saliva samples. Cohorts varied in their inclusion criteria– some admitting only patients who present with fever, some used rash or red eyes, while others admitted patients who presented with fever and/or rash. With the introduction of ZIKV and CHIKV, the PHBDC study chose to adjust the inclusion criteria in order to admit patients who presented with rash, headache and/or fever. Altogether, the longitudinal data of the meta-cohort covers over 18,000 patients (pediatric and adults), in both inpatient and outpatient settings from five countries (Brazil, Colombia, El Salvador, Nicaragua, and Venezuela). Data collection start and end dates vary between cohorts, but ranged from 1998 to the present where the patients have been followed up during different intervals as it can be identified in the table 1 below:

**Table 1.**
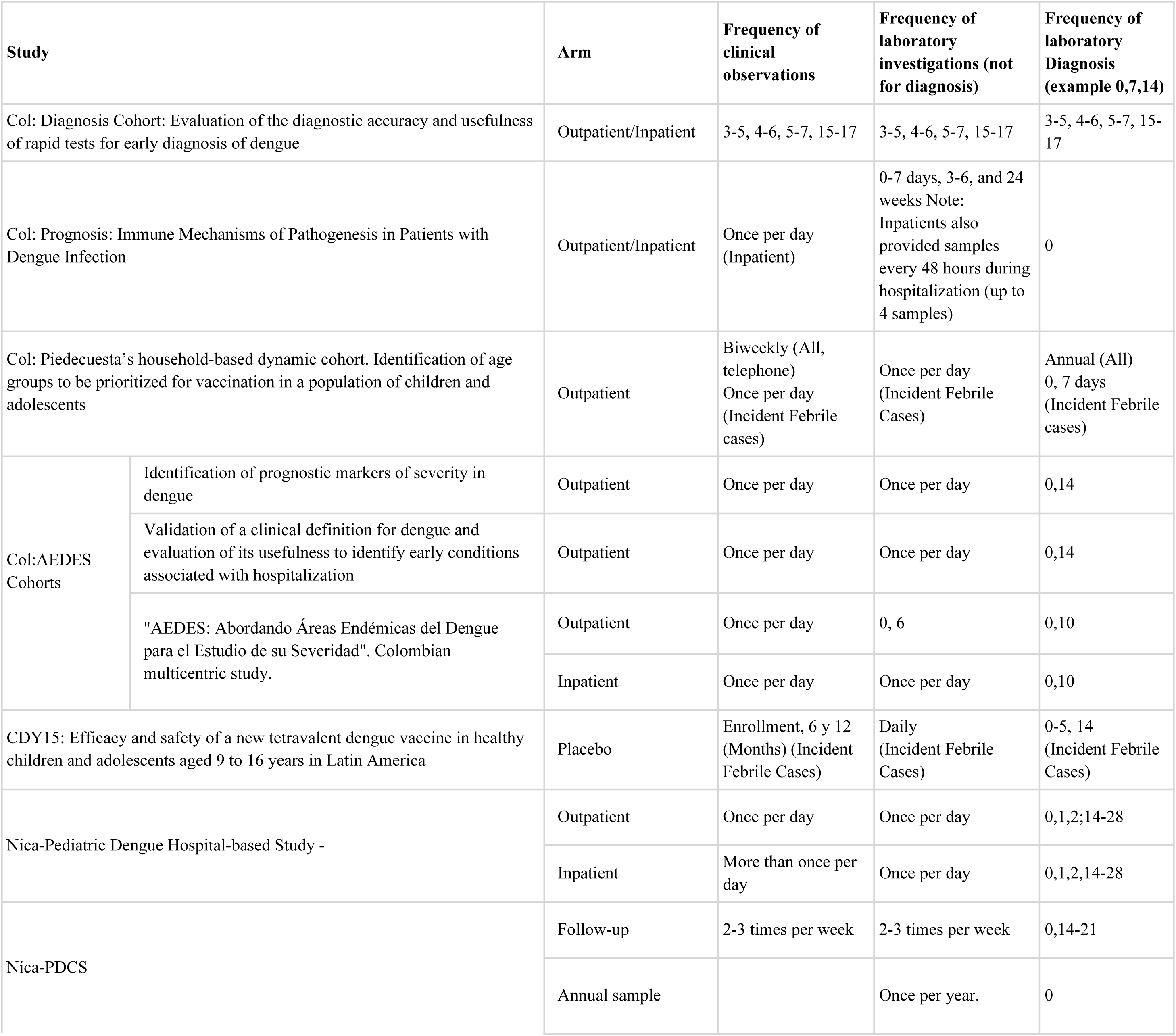
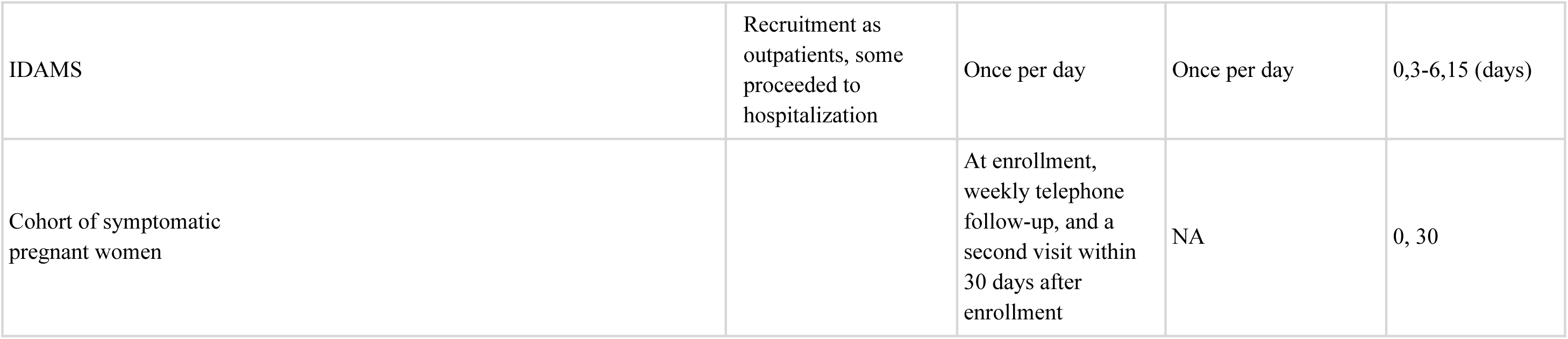
Frequency of follow-up across included cohort studies.

| Study |  | Arm | Frequency of clinical observations | Frequency of laboratory investigations (not for diagnosis) | Frequency of laboratory Diagnosis (example 0,7,14) |
| --- | --- | --- | --- | --- | --- |
| Col: Diagnosis Cohort: Evaluation of the diagnostic accuracy and usefulness of rapid tests for early diagnosis of dengue |  | Outpatient/Inpatient | 3-5, 4-6, 5-7, 15-17 | 3-5, 4-6, 5-7, 15-17 | 3-5, 4-6, 5-7, 15-17 |
| Col: Prognosis: Immune Mechanisms of Pathogenesis in Patients with Dengue Infection |  | Outpatient/Inpatient | Once per day (Inpatient) | 0-7 days, 3-6, and 24 weeks Note: Inpatients also provided samples every 48 hours during hospitalization (up to 4 samples) | 0 |
| Col: Piedecuesta’s household-based dynamic cohort. Identification of age groups to be prioritized for vaccination in a population of children and adolescents |  | Outpatient | Biweekly (All, telephone)<br>Once per day (Incident Febrile cases) | Once per day (Incident Febrile Cases) | Annual (All)<br>0, 7 days (Incident Febrile cases) |
| Col:AEDES Cohorts | Identification of prognostic markers of severity in dengue | Outpatient | Once per day | Once per day | 0,14 |
|  | Validation of a clinical definition for dengue and evaluation of its usefulness to identify early conditions associated with hospitalization | Outpatient | Once per day | Once per day | 0,14 |
|  | "AEDES: Abordando Áreas Endémicas del Dengue para el Estudio de su Severidad". Colombian multicentric study. | Outpatient | Once per day | 0, 6 | 0,10 |
|  |  | Inpatient | Once per day | Once per day | 0,10 |
| CDY15: Efficacy and safety of a new tetravalent dengue vaccine in healthy children and adolescents aged 9 to 16 years in Latin America |  | Placebo | Enrollment, 6 y 12 (Months) (Incident Febrile Cases) | Daily (Incident Febrile Cases) | 0-5, 14 (Incident Febrile Cases) |
| Nica-Pediatric Dengue Hospital-based Study - |  | Outpatient | Once per day | Once per day | 0,1,2,14-28 |
|  |  | Inpatient | More than once per day | Once per day | 0,1,2,14-28 |
| Nica-PDCS |  | Follow-up | 2-3 times per week | 2-3 times per week | 0,14-21 |
|  |  | Annual sample |  | Once per year. | 0 |

|  |  |  |  |  |
| --- | --- | --- | --- | --- |
| IDAMS | Recruitment as outpatients, some proceeded to hospitalization | Once per day | Once per day | 0,3-6,15 (days) |
| Cohort of symptomatic pregnant women |  | At enrollment, weekly telephone follow-up, and a second visit within 30 days after enrollment | NA | 0, 30 |

### Evaluation of the diagnostic accuracy and usefulness of rapid tests for early diagnosis of dengue (Diagnosis Cohort)

The diagnosis cohort, funded by E25bio, Inc., was aimed to determine the diagnostic usefulness of repeated NS1 rapid testing in clinical settings(7). This cohort enrolled and followed patients (≥2 years old) who had both clinically suspected dengue and a positive dengue rapid test (NS1 antigen) at the time of consultation or hospitalization. Participants were followed at 1, 2, and 10 days (convalescence) after recruitment to determine the incidence of dengue complications among confirmed cases. Dengue infection was defined as positive NS1 results (acute sample).

### Immune Mechanisms of Pathogenesis in Patients with Dengue Infection (Prognosis Cohort)

The Prognosis Cohort began with the goal to prospectively validate the predictive accuracy of a pool of transcriptomics intended to predict severity among patients with confirmed DENV infection(8). This study was funded by the U.S. Department of Defense and Colombia’s Centro de Atención y Diagnóstico de Enfermedades Infecciosas (CDI, Colombia). This cohort enrolled patients (≥ 1 year old) with clinically suspected dengue and conducted follow-up at 1, 2, and 10 days (convalescence) after recruitment to determine the incidence of dengue complications among confirmed cases. Participants recruited in outpatient settings were clinically evaluated at enrolment and asked to provide additional blood samples between 3-6 days and 7-10 days if their first sample was obtained up to 2 days and 3-4 days after the onset of fever, respectively. Those who were recruited from inpatient settings underwent daily clinical evaluation, and blood samples were drawn every 48 hours during hospitalization (up to 4 samples). In all participants, additional samples were collected 3-6 and 24 weeks after onset of fever(9,10). Dengue infection was defined as positive PCR results (acute sample).

### Piedecuesta’s Household-Based Dynamic Cohort (PHBDC)

Piedecuesta’s household-based dynamic cohort sought to estimate age-specific dengue seroprevalence and identify age groups to be prioritized for vaccination among children and adolescents. This study was funded by the Colombian Science Ministry, Minciencias. PHBDC was a population-based cross-sectional study which began in 2014 and enrolled and evaluated healthy children and adults (15%) from Piedecuesta (a mid-size city with endemic DENV). The aim of the study was to estimate age-specific dengue seroprevalence in order to identify age groups to be prioritized for vaccination among children and adolescents. Based on the results of this seroprevalence study, a household-based dynamic cohort was initiated in 2015 with the aim of estimating the age-specific incidence of dengue in Piedecuesta (N=2,730). This cohort enrolled children (2-15 years old) and adults within the same household. The cohort followed up with participants on a biweekly basis through telephone contact to identify incident febrile cases. Cases were identified through clinical evaluation, and blood samples studied using Luminex ArboMIA to determine etiological diagnosis (annual [2016–2017] cumulative incidence: 6.0%). Additionally, this cohort conducted annual visits to the participants’ residences to collect blood samples to determine dengue seroconversion (losses to follow-up: 6.5% and 3.2% during the first and second year, respectively), a strategy that allowed for the estimate of attack rates to be calculated for chikungunya (22%) and ZIKV (34%) during the outbreaks of 2015 and 2016, respectively(11).

### Abordando Áreas Endémicas del Dengue para el Estudio de su Severidad (AEDES) Cohorts

The AEDES cohorts’ data is an amalgamation of three independent studies, which were assembled with the aim of developing and validating diagnostic and prognostic algorithms for DENV funded by the same national agency above, Minciencias. The first two cohorts were initiated and conducted during endemic periods (2003-2004, n=500; and 2006-2008, N=705) and a third one during an epidemic (2009-2011, N=2,004). These studies shared similar enrolment protocols: febrile patients with clinically suspected dengue were recruited at the point of care. Whereas the first two cohorts were conducted in outpatient settings of Bucaramanga, the third (the AEDES cohort– see Table 2 for clarification) was a multicenter study that enrolled and followed individuals in both outpatient (Bucaramanga, Barranquilla, and Palmira) and inpatient (Bucaramanga, Cali, Neiva, and Palmira) settings. Patients in the first two cohorts came in for follow-up visits one to seven days via clinical and laboratory assessments (median follow-up: 4 and 3 days, respectively), with a convalescent blood sample taken approximately 2 weeks after disease onset. The third study which enrolled participants from outpatient and inpatient settings had median follow-up times of 3 and 2 days, respectively. Dengue infection was defined as an ELISA IgM/IgG seroconversion or four-fold increase in titers in paired samples or virus isolation (acute sample) in the first two cohorts, and as an ELISA IgM/IgG seroconversion or four-fold increase in paired samples, or positive ELISA NS1 or PCR, or viral isolation (acute sample) in AEDES(12).

**Table 2:**
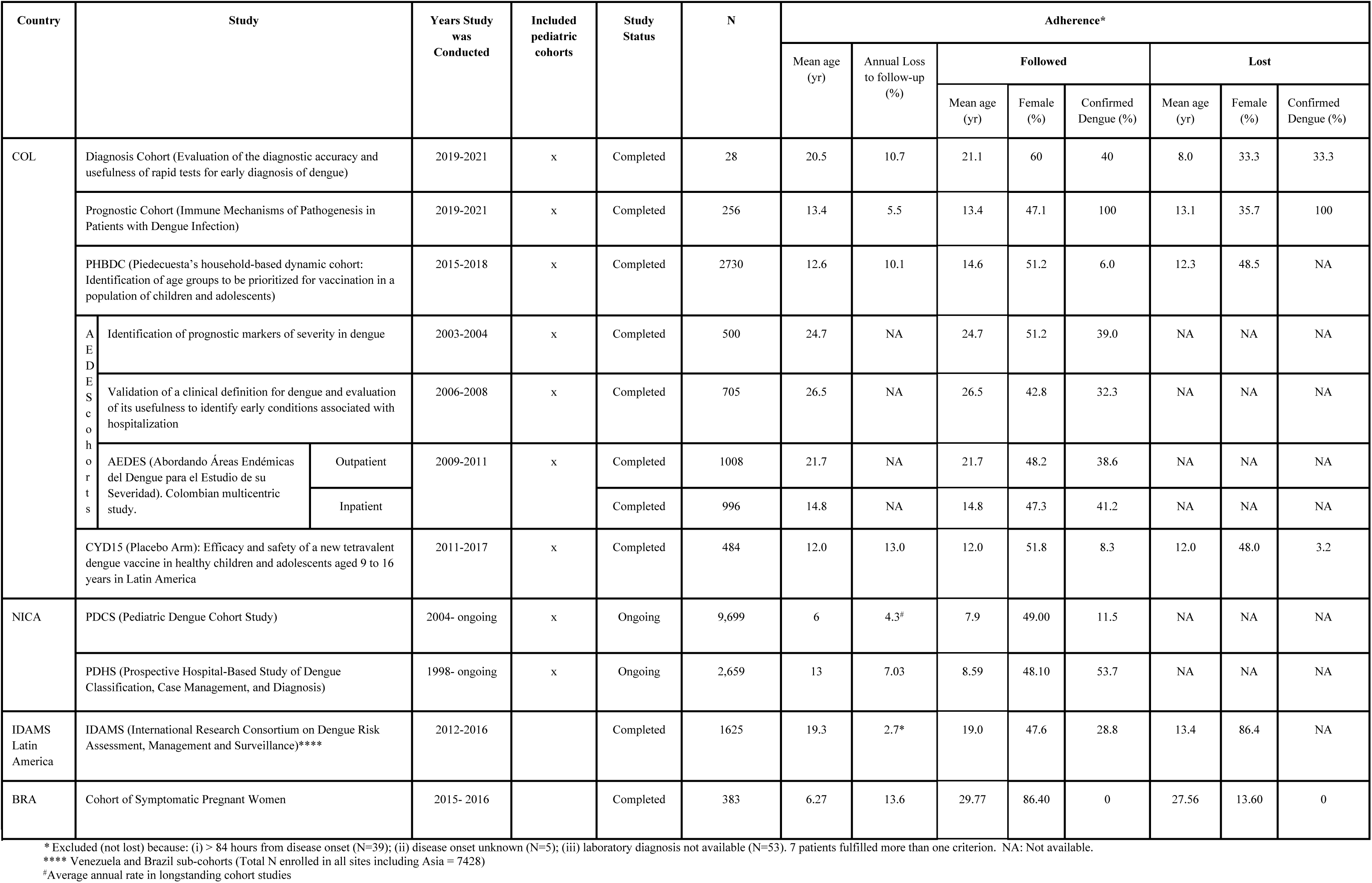
Summary of studies in the Meta-Cohort.

**Table 3.**
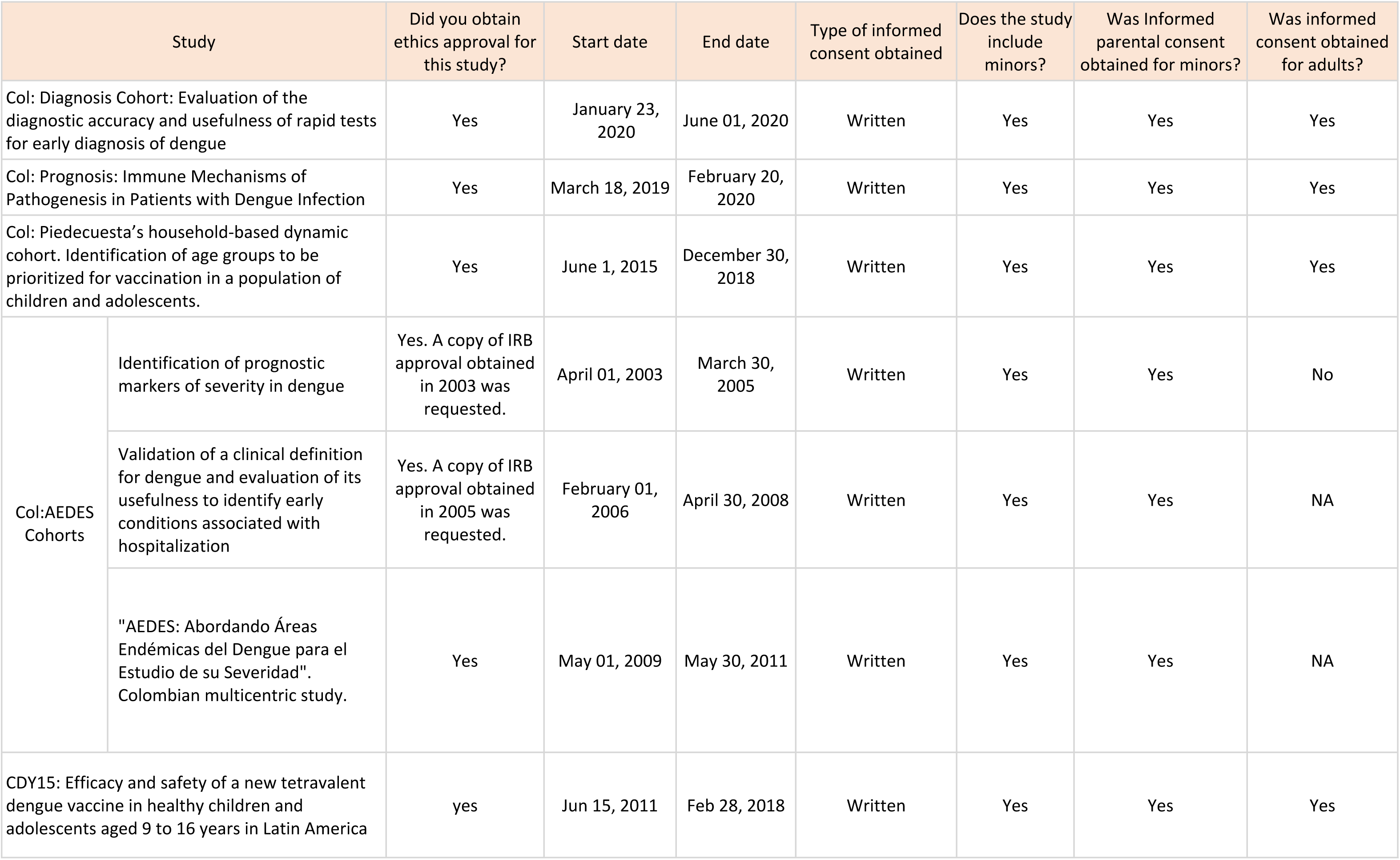
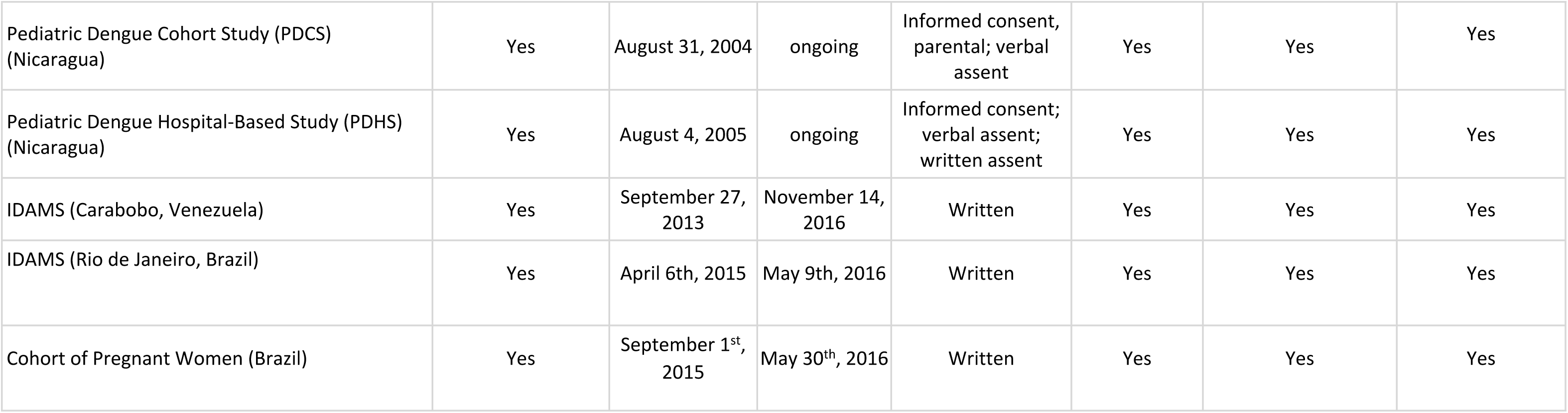
Summary of ethics approvals and consent among patients.

| Study |  | Did you obtain ethics approval for this study? | Start date | End date | Type of informed consent obtained | Does the study include minors? | Was Informed parental consent obtained for minors? | Was informed consent obtained for adults? |
| --- | --- | --- | --- | --- | --- | --- | --- | --- |
| Col: Diagnosis Cohort: Evaluation of the diagnostic accuracy and usefulness of rapid tests for early diagnosis of dengue |  | Yes | January 23, 2020 | June 01, 2020 | Written | Yes | Yes | Yes |
| Col: Prognosis: Immune Mechanisms of Pathogenesis in Patients with Dengue Infection |  | Yes | March 18, 2019 | February 20, 2020 | Written | Yes | Yes | Yes |
| Col: Piedecuesta's household-based dynamic cohort. Identification of age groups to be prioritized for vaccination in a population of children and adolescents. |  | Yes | June 1, 2015 | December 30, 2018 | Written | Yes | Yes | Yes |
| Col:AEDES Cohorts | Identification of prognostic markers of severity in dengue | Yes. A copy of IRB approval obtained in 2003 was requested. | April 01, 2003 | March 30, 2005 | Written | Yes | Yes | No |
|  | Validation of a clinical definition for dengue and evaluation of its usefulness to identify early conditions associated with hospitalization | Yes. A copy of IRB approval obtained in 2005 was requested. | February 01, 2006 | April 30, 2008 | Written | Yes | Yes | NA |
|  | "AEDES: Abordando Áreas Endémicas del Dengue para el Estudio de su Severidad". Colombian multicentric study. | Yes | May 01, 2009 | May 30, 2011 | Written | Yes | Yes | NA |
| CDY15: Efficacy and safety of a new tetravalent dengue vaccine in healthy children and adolescents aged 9 to 16 years in Latin America |  | yes | Jun 15, 2011 | Feb 28, 2018 | Written | Yes | Yes | Yes |

|  |  |  |  |  |  |  |  |
| --- | --- | --- | --- | --- | --- | --- | --- |
| Pediatric Dengue Cohort Study (PDCS)<br>(Nicaragua) | Yes | August 31, 2004 | ongoing | Informed consent,<br>parental; verbal<br>assent | Yes | Yes | Yes |
| Pediatric Dengue Hospital-Based Study (PDHS)<br>(Nicaragua) | Yes | August 4, 2005 | ongoing | Informed consent;<br>verbal assent;<br>written assent | Yes | Yes | Yes |
| IDAMS (Carabobo, Venezuela) | Yes | September 27,<br>2013 | November 14,<br>2016 | Written | Yes | Yes | Yes |
| IDAMS (Rio de Janeiro, Brazil) | Yes | April 6th, 2015 | May 9th, 2016 | Written | Yes | Yes | Yes |
| Cohort of Pregnant Women (Brazil) | Yes | September 1<sup>st</sup>,<br>2015 | May 30<sup>th</sup>, 2016 | Written | Yes | Yes | Yes |

### Efficacy and Safety of a Novel Tetravalent Dengue Vaccine in Healthy Children and Adolescents Aged 9 to 16 years in Latin America - placebo arm (CYD15)

Participants in the CYD15 were healthy children and adolescents (9-16 years old) who were recruited from a placebo arm of a randomized control trial funded by Sanofi Pasteur and had the primary goal of evaluating the efficacy of the chimeric yellow fever–dengue–tetravalent (CYF-DTV) dengue vaccine. The CYD15 study was a multicenter, placebo-controlled, randomized, observer-blind phase III DENV vaccine efficacy clinical trial, examining the efficacy of a vaccine to prevent symptomatic virologically-confirmed dengue cases in infants. Participants were randomly assigned, in a 2:1 ratio, to receive three doses of the Recombinant, live, attenuated, tetravalent dengue vaccine, CYD-TDV, (treatment group) or placebo (0.9% sodium chloride; control group) within 1 year. Participants included in this meta-cohort were healthy children and adolescents between 9-16 years old from the placebo arm living in Bucaramanga, Colombia. Volunteers were invited to participate through contacts with schools in the metropolitan area, referred by relatives of participants, or recruited by community leaders. Participants were followed through biweekly phone calls. In case of identifying any febrile episode, participants were asked to provide blood samples to perform virological confirmation and ELISA (NS1, IgM/IgG) testing for dengue infection as well as hemogram and hepatic function tests. Additional ELISA (IgM/IgG), hemogram and hepatic function tests were repeated in a convalescent sample collected up to 21 days after fever’s onset(13).

### Pediatric Dengue Cohort Study (PDCS)

The PDCS was established as a community-based cohort in District II of Managua, Nicaragua in 2004. The cohort was initially established to study DENV transmission and to characterize symptoms and disease spectrum. It has since evolved to study the virologic and immunologic determinants of response to sequential DENV and ZIKV infections, epidemiological risk factors for infection and disease, and expanded to include other arboviruses, including CHIKV(14,15). The cohort has been funded by a variety of sources, including the US National Institutes of Health (NIH) National Institute of Allergy and Infectious Disease (NIAID), the Pediatric Dengue Vaccine Initiative of the International Vaccine Institute (IVI), the Bill and Melinda Gates Foundation, among others. PDCS is a community-based cohort study which enrolled 2-9-year-olds in August 2004. Participants were originally invited to remain in the study until their 12th birthday, but the restriction increased to 15 in 2007, and 18 in 2019(16). Each year, new two-year old children are enrolled, and additional replacement enrollment is performed as needed in the older age groups to maintain the cohort’s age structure. At any given time, there are roughly 3,800 - 4,100 children actively participating in the PDCS(17,18). With the introduction of chikungunya virus (CHIKV) and ZIKV into Latin America and specifically Nicaragua, CHIKV and ZIKV were added to the PDCS in 2014 and 2015, respectively. Visits are divided into 4 categories (A-D) based on symptomatology(19). Category A cases include fever plus symptoms and signs of suspected dengue (WHO’s case definition). Category B cases are undifferentiated febrile illnesses. Category C cases are fevers with a non-arboviral diagnosis (e.g., influenza, UTI), and category D cases are non-febrile cases. With the introduction of ZIKV, the D case category was divided into two subsets, D cases with ZIKV-like symptoms such as red eyes and rash and D cases without ZIKV-like symptoms(20).

Category A, B, and ZIKV-like D cases are tested for DENV, ZIKV, and CHIKV using RT-PCR and serological testing. Acute (1-4 days post-onset of symptoms) and convalescent (14-21 days) samples are collected from all suspected cases. An additional sample is collected at day 4-6 from RT-PCR-confirmed DENV and ZIKV cases for immunological studies(21). Each year, a healthy blood sample (serum/plasma, and PBMCs prepared from a subset) is collected and used (a) to detect arbovirus infections that may not have been apparent but may have occurred throughout the year and (b) for additional immunological studies. Data on socioeconomic factors, demographics, and medical history are collected at enrollment and are updated annually. During clinical visits, detailed information on symptoms and symptom onset is collected(22).

### Pediatric Dengue Hospital-Based Study (PDHS)

This cohort was founded in 1998 to investigate the clinical, immunological, and viral risk factors for severe DENV, assessing biomarkers, and studying immune responses over time. This study has been supported by the NIAID at NIH through various mechanisms. The Pediatric Hospital-Based Study began in 1998 and enrolls children ages 6 months to 14 years who present to the Hospital Infantil Manuel de Jesus Rivera (HIMJR) with suspected dengue (< 7 days from illness onset)(23). Both in-patient and out-patient suspected cases are eligible for enrollment. Upon enrollment, a complete physical exam is performed, and medical history is collected. Participants are followed throughout the acute phase of their illness and data including vital signs, symptoms, and treatment are recorded daily. Blood samples for complete blood counts, molecular, serological, and virological testing are collected daily for the first three days. An additional convalescent sample is collected 14-21 days after enrollment. A longitudinal arm of this study, for those participants who consent, collects samples 3-, 6-, 12-, and 18-months post-illness for immunological studies. The protocol was amended in 2014 to include CHIKV and again in 2016 to include ZIKV.

### International Research Consortium on Dengue Risk Assessment, Management and Surveillance (IDAMS)

The primary objective of the IDAMS Cohort was to evaluate warning signs and predictors or biomarkers associated with progression to severe dengue in order to facilitate triage efforts. Funding was provided by the EC’s Seventh Framework Program. The IDAMS study (2011-2016) was a prospective multi-center acute febrile illness study conducted in Vietnam, Cambodia, Malaysia, Indonesia, Brazil, Venezuela, and El Salvador. Sites recruited participants who with a history of fever for ⩽72-84 hours (site-dependent), presenting with clinical symptoms suggestive of dengue (in patients >5 years of age). Patients were excluded if a) they presented with severe dengue at enrolment, b) a clinician judged the patient was unlikely to attend daily outpatient follow-up visits, or c) the clinical presentation strongly suggested a diagnosis other than dengue (e.g., pneumonia, otitis, etc.). Only the data from Latin America were considered for the meta-cohort. However, as the data structure is homogenous, other study locations could be added in the future. The study design was described before in detail (24,25). In brief, patients with a history of fever for less than 72-84 hours (site dependent) and suggestive of DENV were recruited in outpatient clinics across the participating sites and followed up daily for a maximum of 6 days or until afebrile for 48h, with a final follow-up visit around day 10-14 of illness. Daily follow-up included physical examination as well as simple laboratory investigations such as full blood count. Dengue infection was defined as confirmed positive by either PCR or ELISA NS1 result in acute sample.

### Cohort of Symptomatic Pregnant Women

In 2012, a prospective cohort for dengue surveillance in mother–infant pairs was established within the Manguinhos, Rio de Janeiro area. In 2015, however, most of these were later identified as ZIKV cases. To identify these ZIKV cases in the Rio de Janeiro population, the pregnancy cohort study was modified to enroll women who presented with a rash at any week of gestation. It was supported by the Department of Science and Technology (Departamento de Ciência e Tecnologia– DECIT) of the Brazilian Ministry of Health (Ministério da Saúde) and funded by the Coordination of the Improvement of Higher Level Personnel (Coordenação de Aperfeiçoamento de Pessoal de Nível Superior-CAPES); the Bill and Melinda Gates Foundation, Grand Challenges Explorations; and the National Institute of Allergy and Infectious Diseases (NIAID) of the National Institutes of Health. Brazil’s Symptomatic Women cohort offered enrollment between September 2015 to September 2016 to pregnant women who attended the acute febrile illness clinic at the Oswaldo Cruz Foundation and who presented with a rash that had developed in the previous 5 days, with or without an associated fever. Laboratory data was collected after enrollment(26). Weekly follow-ups occurred over the phone, and clinical and laboratory follow-up occurred within 30 days of enrollment and were referred for fetal ultrasonography follow-ups at three timepoints: before 20 weeks of gestation, between 20-30 weeks of gestation, and after 30 weeks of gestation. ZIKV infection was defined as positive RT-PCR, in acute samples of blood or urine.

### Retrospective and Prospective Harmonization Efforts of the Meta-Cohort

Individual patient data (IPD) meta-analyses (MA) are considered the gold standard for meta-analyses(27)The strength of conducting IPD-MAs, as opposed to standard meta-analyses using effect estimates, is the opportunity to control for baseline heterogeneity

Knowing that we had a collective wealth of arbovirus data, we initially set out to find a common data dictionary for all cohorts within the ReCoDID consortium. This effort included trying to find common variables across all zika cohorts, then across dengue cohorts, as well as any chikungunya patients. As mentioned in the literature, there has not been a gold standard method for data harmonization, but we eventually found the Maelstrom Guide to Rigorous Retrospective Harmonization (28). The first step of the guide is to define a research objective for harmonization, which allowed us to focus on the dengue studies based on a research question that aimed to identify future flavivirus clinical epidemiology in settings where we can infer the past history of infections. Once we had a focused research question, the next step was implementing a well-defined structure of dimensions (endpoints/confounders/exposure) and domains to tackle from a medical perspective. These dimensions guided us in establishing a set of medical meetings where we identified, prioritized and defined in a semantic manner the variables to include in the master data dictionary as indicated in the methodology used and explained in the Fig 1.

**Fig 1.**
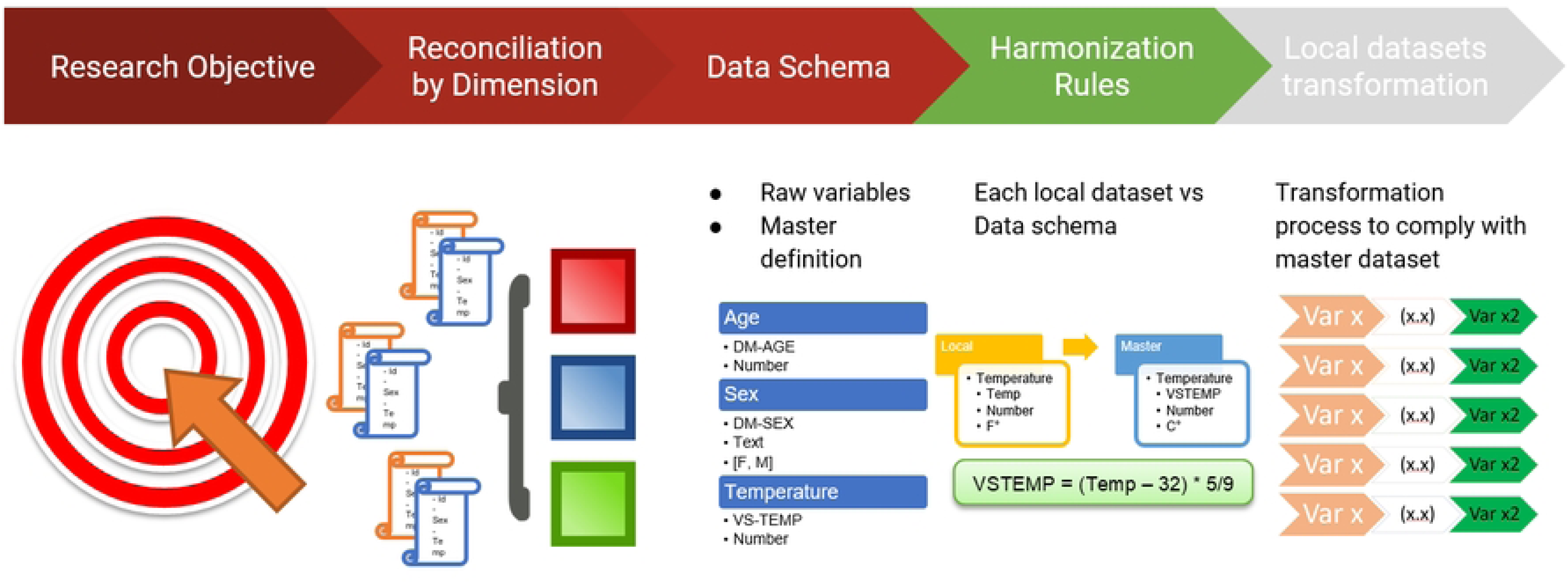
Harmonization process.

Then we compared each study data dictionary against the reconciled master data dictionary establishing a harmonization potential based on a predefined structure of conventions that gave us a complete set of variables with different levels of potential use for harmonization, this can be identified in the Fig 2.

**Fig 2.**
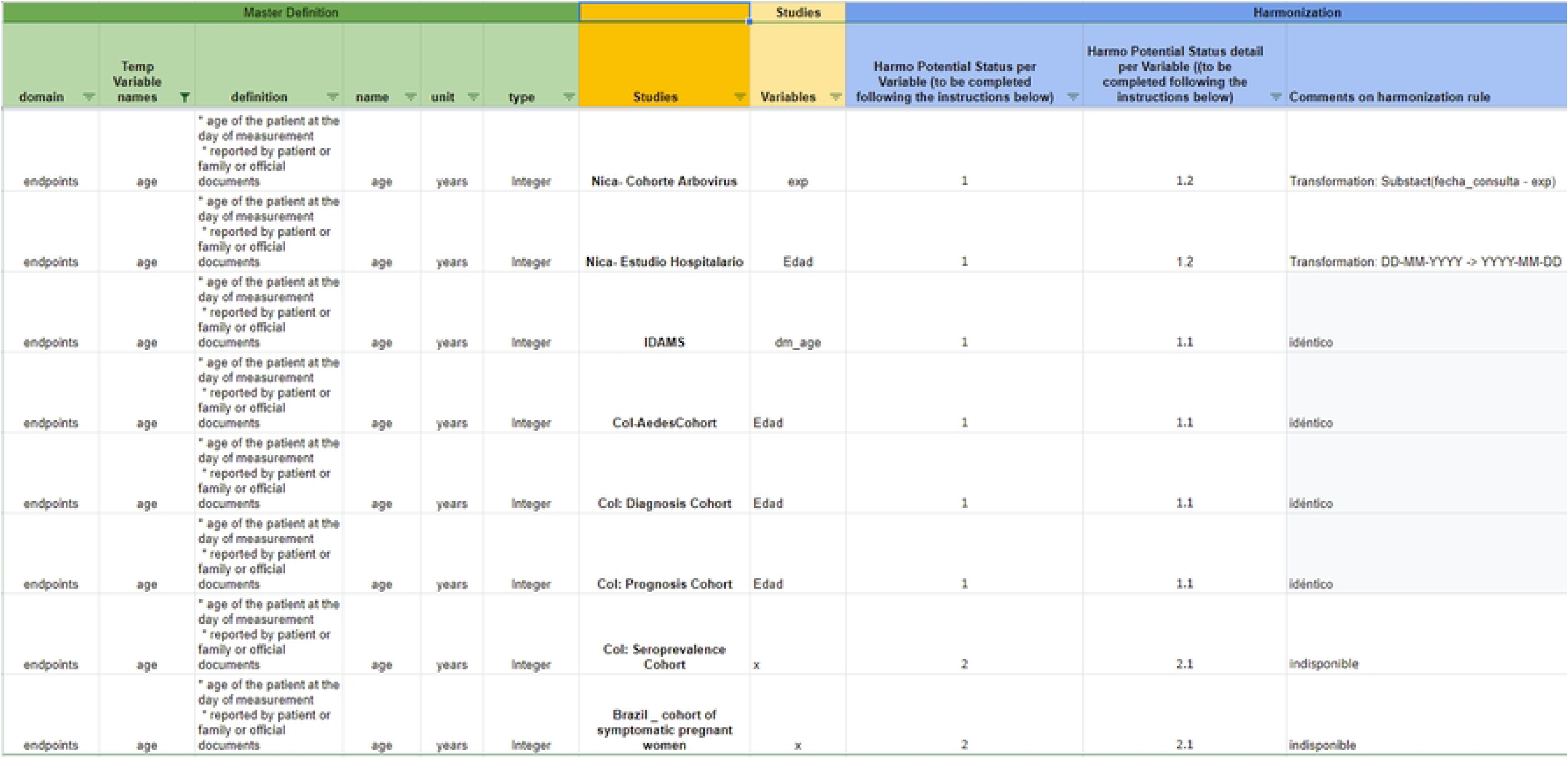
Harmonization potential

Retrospective harmonization is generally resource-intensive (29,30), as we can attest to. In order to facilitate future IPD-MAs, we are in the process of creating a Case Report Form which we then standardized according to the CDISC data standard. The hope is that future cohorts conducting arbovirus research will be able to prospectively implement this CRF, decreasing, or removing entirely, the effort and cost of retrospectively harmonizing data to be pooled in an IPD-MA.

### What has it found? Key findings and publications

Key findings include: 1) low serum 25(OH)D concentrations in patients predicted the progression of dengue fever(31); 2) the highest risk group for severe dengue being patients with preexisting anti-DENV antibody titers; the same study showing a preventative effect in those patients who have a (very) high level of antibody titers(32); and 3) the development, validation, and evaluation of the usefulness of a scale for the prediction of disease severity among confirmed cases of dengue (32%, 39%, and 41% for the three cohorts).

### What are the main strengths and weaknesses of the combined meta-cohort effort?

A technical strength of the Consortium’s efforts is the retrospective harmonization of participant-level data which has been completed according to the Maelstrom group’s recommendations for best practice(28), which outlines steps 0 (define the research question) to 5 (disseminate and preserve final harmonization products). Maelstrom Research guidelines’ Step 0 is why the ReCoDID Consortium’s data outlined in this profile is primarily focused on dengue-related outcomes despite collecting data related to viruses. Another strength is the use of CDISC/SDTM standards as reference for the definition of the core variables harmonized (33), which enables interoperability between the meta-cohort and other studies that apply CDISC SDTM. The harmonization (processing data collected by studies und a common variable format), or standardization (use of a data standard (CDISC, e.g.) to define the core variables format to be generated, required to conduct individual-level meta-analyses can be extremely resource-demanding. Going forward, and in order to minimize this burden and improve individual-level patient data meta-analyses (IPD-Mas), the ReCoDID Consortium recommends the creation of a standard case report form (CRF) for acute viral syndrome (arbovirus) research, which includes the features of the overlapping clinical syndromes associated with the most important arboviruses (e.g., DENV, ZIKV, CHIKV, etc.), to be used by all partner studies in the future. Potentially, this CRF will include modules for different severe disease manifestations that can be adapted to the local situation (i.e., bleeding module, neurological module, liver pathology module).

Another strength of the combined cohort is that it covers different countries and partner sites across Latin America - each of which experienced slightly different histories of DENV serotypes, CHIKV, and ZIKV infections over the last decades. The resulting ‘experiment of nature’ represents a population immune landscape that we now would like to prospectively follow with future cohorts, considering the potential of immunological interaction between related flaviviruses (e.g., DENV1-4 and ZIKV). In Fig 3 and Fig 4, created using R (4.0.5)(34), we present DENV, ZIKV, and CHIKV activity over time in the respective countries and partner sites. This combined cohort is a first step towards the direction of a multicentric arbovirus cohort, which needs to be complemented with advanced technology assessing immunological history, additional investments in future harmonization, and standardization.

**Fig 3.**
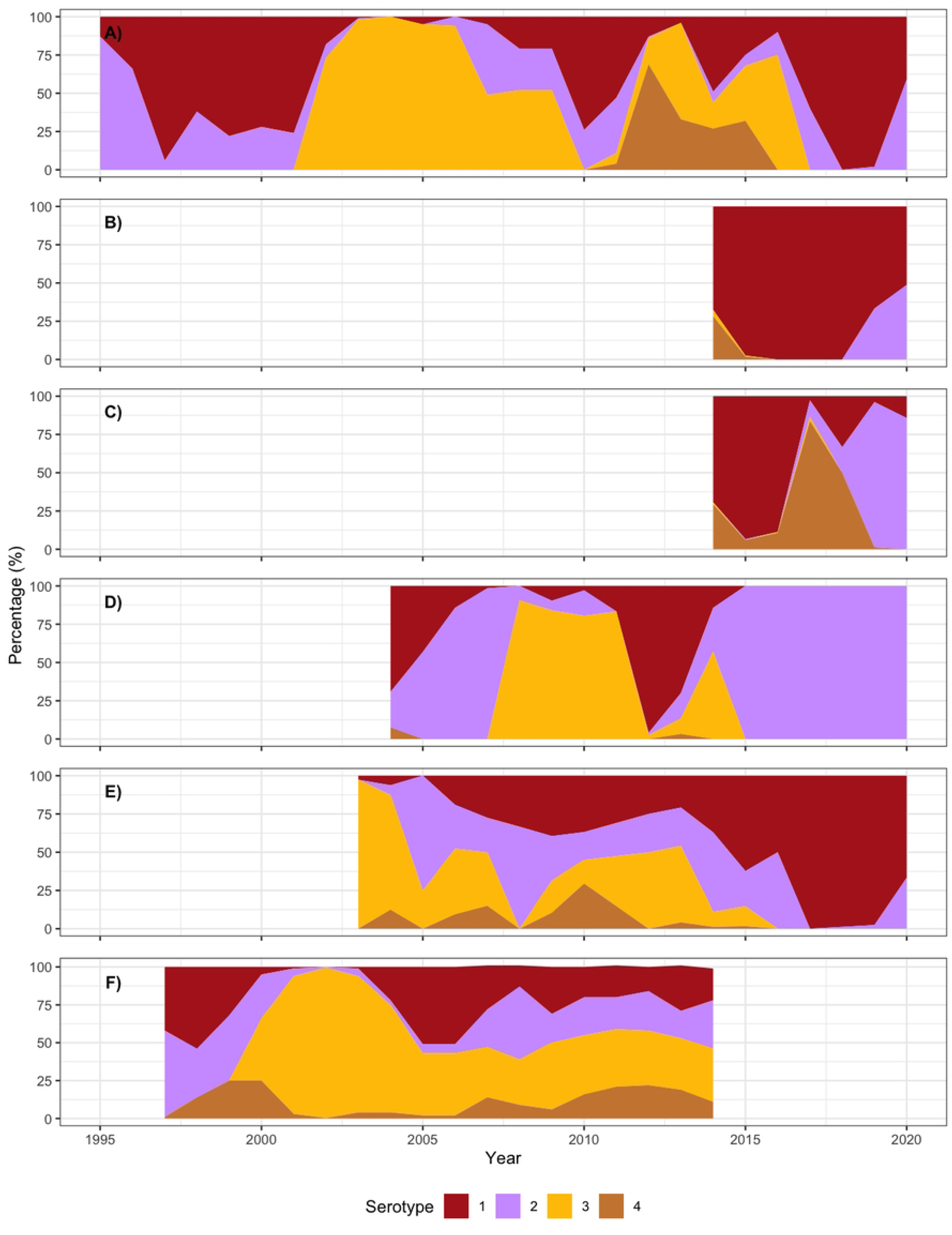
DENV serotype distribution over time. a) Pernambuco (Brazil) * b) Ceará (Brazil) Δ c) Rio de Janeiro (Brazil) Δ d) Nicaragua § e) Colombia ∞ f) Venezuela # * Data of annual DENV serotypes from PE, provided by: the Central Laboratory of Pernambuco (Laboratório Central de Pernambuco-LACEN PE) Δ Data of annual DENV serotypes from CE and RJ, retrieved from the national online database of the Brazilian Ministry of Health (https://datasus.saude.gov.br/informacoes-de-saude-tabnet/, accessed on 11/8/21 to 11/11/21 § PDCS arboviral case data (Oct 2004 - Mar 2021). Image one contains yearly level data of PCR confirmed Dengue cases, and image two contains DENV, CHIKV and ZIKV confirmed symptomatic infections on a monthly basis. ∞ This data was derived from the Colombian National Institute of Health. http://portalsivigila.ins.gov.co/Paginas/Vigilancia-Rutinaria.aspx) # Dengue incidence data (1997-2014), Data from: the National Surveillance System of the Venezuelan mandatory notification diseases,the Ministry of Health (http://www.mpps.gob.ve). Data on the proportion of dengue cases per serotype in Aragua, provided by: the Laboratorio Regional de Diagnóstico e Investigación del Dengue y otras Enfermedades Virales (LARDIDEV), Corporación de Salud Aragua, Maracay, Venezuela, and published in Lizarazo Forero, EF (2019). Epidemiology, genetic diversity and clinical manifestations of arboviral diseases in Venezuela. PhD Thesis. University of Groningen, Groningen, the Netherlands. https://doi.org/10.33612/diss.108089934

**Fig 4.**
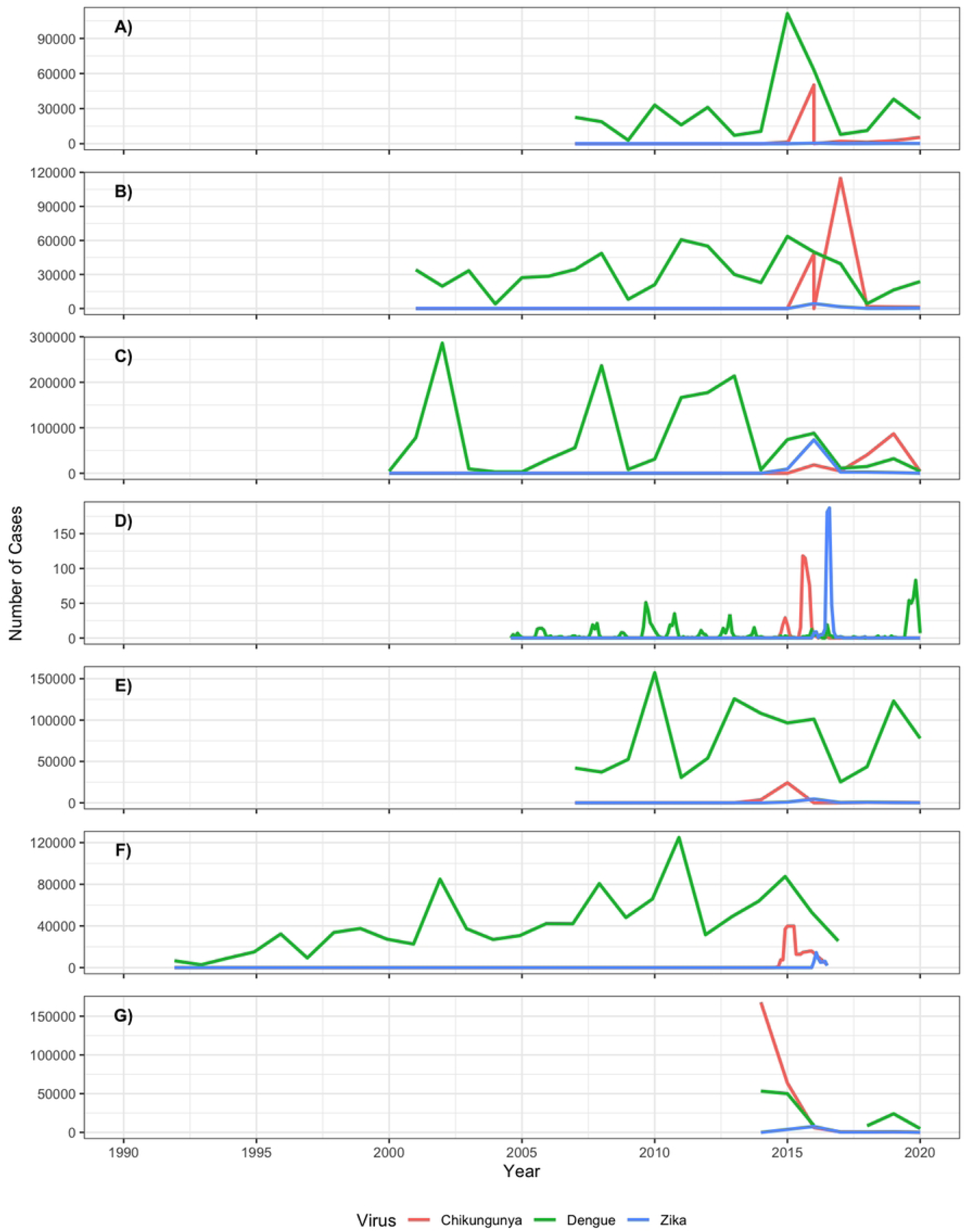
Reported number of cases for DENV, ZIKV and CHIKV over time a) Pernambuco (Brazil) ∐, ∑,+ b) Ceará (Brazil) ∐, + c) Ceará (Brazil) ∐, + d) Nicaragua § e) Colombia ∞ f) Venezuela ** g) El Salvador ∐ Data of annual DENV, ZIKV and CHIKV, reported cases in the states of Ceará (CE), Pernambuco (PE) and Rio de Janeiro (RJ), retrieved from: the national online database of the Brazilian Ministry of Health (https://datasus.saude.gov.br/informacoes-de-saude-tabnet/, accessed on 11/8/21 to 11/11/21), except for CHIKV in 2015-2016 in CE and PE (see below). Data of CHIKV from CE and PE (2015-2016), taken from: Epidemiological Bulletins (EBs) published by the Brazilian Ministry of Health. ∑ Data for ZIKV from PE (2015), may be subject to reporting bias. At the time, ZIKV was largely unknown and therefore ZIKV cases might have been classified/notified as dengue cases, de Brito et al. (2016), Data from: Rev Soc Bras Med Trop 49(5):553-558, September-October, 2016 doi:10.1590/0037-8682-0245-2016.. + Data of CHIKV from CE and PE (2015-2016), Taken from; Epidemiological Bulletins (EBs) published by the Brazilian Ministry of Health. § PDCS arboviral case data (Oct 2004 - Mar 2021). Image one contains yearly level data of PCR confirmed Dengue cases, and image two contains DENV, CHIKV and ZIKV confirmed symptomatic infections on a monthly basis. ∞ This data was derived from the Colombian National Institute of Health. http://portalsivigila.ins.gov.co/Paginas/Vigilancia-Rutinaria.aspx) ** a) Vincenti-Gonzalez MF, A Tami, EF Lizarazo, ME Grillet (2018), Data from: ENSO-driven climate variability promotes periodic major outbreaks of dengue in Venezuela. Scientific Reports, Apr 10;8(1):5727. doi: 10.1038/s41598-018-24003-z b) Data derived from: Boletines Epidemiológicos de Venezuela, currently found at https://www.ovsalud.org/publicaciones/documentos-oficiales/. c) Data from: Venezuela National EPI-12 notifications, weeks 1-29, (2016). d) PAHO/WHO Epidemiological weekly bulletins at https://www.paho.org/en/documents

### Can I access the data? Where can I find out more?

Interested parties will be able to access data dictionaries that include information on variables across the datasets via <u>BioStudies</u>(35). After consultation with each cohort, linked harmonized and curated human cohort data (CE and HDL) is planned to be made accessible through the <u>European</u> <u>Genome-phenome Archive (EGA)</u>(36) platform to Data Users after their requests are evaluated by the ReCoDID Data Access Committee (DAC). Researchers will be able to openly access the descriptive cohort metadata related to the meta-cohort via the EMBL-EBI <u>Cohort Browser</u>(37). Cohorts sharing CE and HDLdata within the ReCoDID Platform are responsible for obtaining regulatory and ethical approvals at the local, or, in the case of Brazil, national, level for data sharing. Where cohorts’ original informed consent forms did not include broad consent for future use of data, ReCoDID worked with cohorts to apply for a waiver of consent. The waivers of consent submitted to the Commission of Ethics in Research, CONEP, Brazil’s national ethics regulatory authority. The waivers of consent have been in process for one year and we expect we will need two years to complete the CONEP review.. Samples will not be shared; however, the associated viral sequencing data can be shared and accessed openly at the <u>European Nucleotide</u> <u>Archive (ENA)</u>(38). Any further associated and shared data types can be shared and retrieved from the relevant <u>EMBL-EBI resources</u>(39).

The assessment of political, ethical, administrative, regulatory, and legal (PEARL) issues related to data sharing guided the implementation and activities of ReCoDID to promote ethical data governance and sharing that carefully considers the perspective and context of Low and Middle Income Countries (LMICs). Empirical research, bibliography reviews, and stakeholders’ consultations guided ReCoDID members’ activities and informed the development of the ReCoDID Data Governance Framework (DGF). The ReCoDID DGF is a high-level normative, organizational, and technical document that describes the goals and principles by which the ReCoDID functions, among them the FAIR (findable, accessible, interoperable, and reusable) data principles. The ReCoDID DGF also implements international standards and best practices for data sharing to promote the public interest and advancement of science. It further describes how the ReCoDID Platform functions and complies with data protection and privacy legislation, mainly the European Union (EU) General Data Protection Regulation (GDPR), and how it relates to other countries’ legislations considering international data transfers. The ReCoDID DGF is centred on protecting the rights and interests of Data Subjects (patients and research participants) and different stakeholders that participate in the biomedical innovation ecosystem, including researchers and information technology (IT) professionals that engage in biomedical research and develop the infrastructure and tools that enable health data platforms.

ReCoDID acknowledges the challenges to promoting a biomedical innovation system that is transparent, equitable, and participatory, that incorporates LMICs’ context-specific perspectives. This is deeply rooted in identifying and overcoming the PEARL barriers related to data sharing mentioned above, as well as the enablers and different strategies to guarantee and implement ethical data sharing and governance. Addressing these PEARL barriers requires creating discussion forums, building research networks, and promoting best practices among academic communities that call for the de-colonization of global health practices and challenge power structures that support and perpetuate them. Therefore, the ReCoDID DGF and ReCoDID Intellectual Property and Open Science Policy incorporate these elements and include issues related to benefit sharing, authorship, attribution, and recognition, as well as the mechanism to implement them.

For more information, please contact, with reference to the ReCoDID Project. To share associated human cohort data, please contact the University of Heidelberg.

## Funding

ReCoDID is supported by the European Union’s Horizon 2020 Research and Innovation Programme under Grant Agreement No. 825746 and the Canadian Institutes of Health Research, Institute of Genetics (CIHR-IG) under Grant Agreement N.01886-000. The funders had no role in study design, data collection and analysis, decision to publish, or preparation of the manuscript.

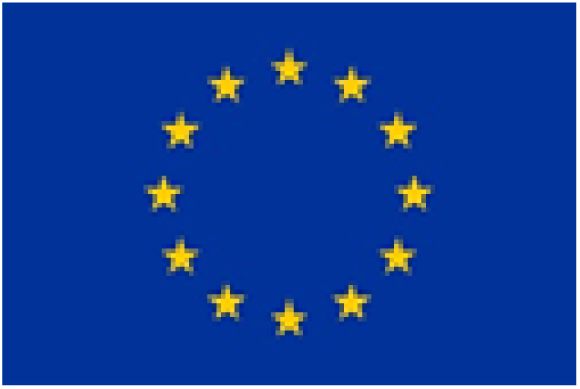

## Data Availability

Yes, data will be available. We describe this in a particular section of our manuscript.

https://ega-archive.org/

https://www.ebi.ac.uk/ena/browser/home

https://www.pathogensportal.org/

https://www.ebi.ac.uk/biostudies/

## Acknowledgements

This work was supported by the ReCoDID Arbovirus harmonization study group:

***Janet Achieng*** *(Heidelberg Institute of Global Health (HIGH), Heidelberg University Hospital, Heidelberg, Germany)*

***Sonia Arguello*** *(Oswaldo Cruz Foundation (Fiocruz), Rio de Janeiro, Brasil)*

***Gabriela Maria Marón Alfaro*** *(Hospital Nacional de Niños Benjamín Bloom, San Salvador, El Salvador)*

***Till Bärnighausen*** *(Heidelberg Institute of Global Health (HIGH), Heidelberg University Hospital, Heidelberg, Germany)*

***Bruno Souza Benevides*** *(University of the State of Ceara, Fortaleza, Brazil)*

***Sarah Bethencourt*** *(Departamento de Estudios Clínicos, Facultad de Ciencias de la Salud, Universidad de Carabobo, Valencia, Venezuela)*

***Andrea Caprara*** *(Departamento de Estudios Clínicos, Facultad de Ciencias de la Salud, Universidad de Carabobo, Valencia, Venezuela)*

***Priscilla M.D.G. César*** *(McMaster University, Institute on Ethics & Policy for Innovation (IEPI))*

***Guy Cochrane*** *(European Molecular Biology Laboratory, European Bioinformatics Institute, Wellcome Genome Campus, Hinxton, Cambridge CB10 1SD, UK)*

***Monika Consuegra*** *(Centro de Atención y Diagnóstico de Enfermedades Infecciosas, Bucaramanga, Colombia)*

***Fabio Otero*** *(Centro de Atención y Diagnóstico de Enfermedades Infecciosas, Bucaramanga, Colombia)*

***Kerstin Rosenberger*** *(Heidelberg Institute of Global Health (HIGH), Heidelberg University Hospital, Heidelberg, Germany)*

***Anna M. Gajewski*** *(Sustainable Sciences Institute, Managua, Nicaragua)*

***Nadim Rahman*** *(European Molecular Biology Laboratory, European Bioinformatics Institute, Wellcome Genome Campus, Hinxton, Cambridge CB10 1SD, UK)*

***Bladimir Cruz*** *(Hospital Nacional de Niños Benjamín Bloom, San Salvador, El Salvador)*

***Guillermo Barahona Escobar*** *(Hospital Nacional de Niños Benjamín Bloom, San Salvador, El Salvador)*

***Maria I. Estupiñán*** *(Centro de Atención y Diagnóstico de Enfermedades Infecciosas, Bucaramanga, Colombia)*

***Isabel Fortier*** *(Research Institute, McGill University Health Centre, Montreal, Canada)*

***Rosa Margarita Gélvez Ramírez*** *(Centro de Atención y Diagnóstico de Enfermedades Infecciosas, Bucaramanga, Colombia)*

***María Fernanda Vincenti Gonzalez*** *(University of Groningen, University Medical Center Groningen, Department of Medical Microbiology and Infection Prevention, Groningen, The Netherlands)*

***Peter W. Harrison*** *(European Molecular Biology Laboratory, European Bioinformatics Institute, Wellcome Genome Campus, Hinxton, Cambridge CB10 1SD, UK)*

***Suran Jayathilaka*** *(European Molecular Biology Laboratory, European Bioinformatics Institute, Wellcome Genome Campus, Hinxton, Cambridge CB10 1SD, UK)*

***Guillermina Kuan*** *(Sustainable Sciences Institute, Managua, Nicaragua)*

***Manish Kumar*** *(European Molecular Biology Laboratory, European Bioinformatics Institute, Wellcome Genome Campus, Hinxton, Cambridge CB10 1SD, UK)*

***Luz Marina Leegstra*** *(Heidelberg Institute of Global Health (HIGH), Heidelberg University Hospital, Heidelberg, Germany)*

***Erley Ferlipe Lizarazo*** *(University of Groningen, University Medical Center Groningen, Department of Medical Microbiology and Infection Prevention, Groningen, The Netherlands)*

***Luigi Marongiu*** *(Heidelberg Institute of Global Health (HIGH), Heidelberg University Hospital, Heidelberg, Germany)*

***Ágnes Molnár*** *(Heidelberg Institute of Global Health (HIGH), Heidelberg University Hospital, Heidelberg, Germany)*

***Cesar Narvaez*** *(Sustainable Sciences Institute, Managua, Nicaragua)*

***Federico Narvaez*** *(Sustainable Sciences Institute, Managua, Nicaragua)*

***Sergio Ojeda*** *(Sustainable Sciences Institute, Managua, Nicaragua)*

***Gabriele Rinck*** *(European Molecular Biology Laboratory, European Bioinformatics Institute, Wellcome Genome Campus, Hinxton, Cambridge CB10 1SD, UK)*

***Ernesto Pleités Sandoval*** *(Hospital Nacional de Niños Benjamín Bloom, San Salvador, El Salvador)*

***Frank Tobian*** *(Heidelberg Institute of Global Health (HIGH), Heidelberg University Hospital, Heidelberg, Germany)*

***José Victor Zambrana*** *(Sustainable Sciences Institute, Managua, Nicaragua)*

*16. **Luana Damasceno** (Oswaldo Cruz Foundation (Fiocruz), Rio de Janeiro, Brasil)*

